# Impact of School Reopening on Pandemic Spread: A Case Study using an Agent-Based Model for COVID-19

**DOI:** 10.1101/2021.03.13.21253485

**Authors:** Hanisha Tatapudi, Tapas K. Das

**Author notes:** Corresponding author: Hanisha Tatapudi – corresponding author., Tapas K. Das.

## Abstract

This article examines the impact of partial/full reopening of school/college campuses on the spread of a pandemic using COVID-19 as a case study. The study uses an agent-based simulation model that replicates community spread in an urban region of U.S.A. via daily social mixing of susceptible and infected individuals. Data representing population demographics, SARS-CoV-2 epidemiology, and social interventions guides the model’s behavior, which is calibrated and validated using data reported by the government. The model indicates a modest but significant increase (8.15 %) in the total number of reported cases in the region for a complete (100%) reopening compared to keeping schools and colleges fully virtual. For partial returns of 75% and 50%, the percent increases in the number of reported cases are shown to be small (2.87% and 1.26%, respectively) and statistically insignificant. The AB model also predicts that relaxing the stringency of the school safety protocol for sanitizing, use of mask, social distancing, testing, and quarantining and thus allowing the school transmission coefficient to double may result in a small increase in the number of reported infected cases (2.14%). Hence for pandemic outbreaks from viruses with similar characteristics as for SARS-CoV-2, keeping the schools and colleges open with a modest campus safety protocol and in-person attendance below a certain threshold may be advisable.

## INTRODUCTION

When the COVID-19 pandemic hit the U.S. in late February of 2020, policy makers across the nation implemented a lockdown by closing non-essential businesses and switching to virtual operation for schools/colleges and some of the essential workplaces. By August 2020, as several regions in the U.S. saw a decline in the number of new cases, the school districts decided to either partially or fully reopen school/college campuses for students to return. We developed a plan for a model-based study of the impact school/college reopening had on the community by comparing the increase in the number of infected cases between continued virtual operation and various levels of reopening (50%, 75%, and 100%). Since many adverse events took place soon after reopening of schools/colleges in September of 2020, such as setting in of the winter weather prompting people to be indoors, U.S. presidential election rallies in September and October, Thanksgiving holidays in the last week of November, and Christmas holidays, the total number of infections increased immensely in the last months of 2020. Hence, a key question that we examined is what portion of the increased cases was contributed by the reopening of schools/colleges. This paper presents our findings and conclusions, which we believe will be useful for decision makers in potential future pandemic outbreaks of similar (SARS) virus types.

To aid our investigation, we developed a comprehensive AB simulation model that mimics pandemic spread using COVID-19 outbreak in an urban outbreak region as a case study; the region we focused on is Miami-Dade County of Florida, USA with 2.8 million population. The AB model yields estimate for age-stratified numbers of actual infections, reported cases, hospitalized, and dead. The model was calibrated and validated using publicly available data from the region until end of September 2020 as schools reopened on September 30. We froze the model parameters to their calibrated values till end September and ran the simulation till the end of December 2020 with school reopening as the only major new event. We examined various reopening scenarios (e.g., 50%, 75%, and 100% return to campus) and different estimated values of transmission coefficients at schools and colleges. Transmission coefficients were assumed to be 1.5x, 2x, 2.5x, and 3x the estimated transmission coefficient for essential workplaces, where control measures are easier to implement and maintain. Results from different scenarios were compared with those for the respective base cases (0% student return and 1.5x school transmission coefficient).

For pandemics before COVID-19, school closure was considered an important and impactful social intervention [1]. Several studies examined the impact of school closure strategies [1-6]. However, the strategies evaluated in many of these studies were implemented for a limited time window, after which a complete reopening occurred with minor variations. In contrast, COVID-19 has persisted for most of 2020 and is continuing into 2021, and thus, in addition to school closure policies, there is also a need to understand school reopening policies. Another distinctive difference between COVID-19 and the past pandemics is that the latter have mostly been caused by influenza viruses, which affects children and older adults more than the other age groups, whereas COVID-19 affects older adults more.

Several studies have evaluated the impact of school closure and reopening for COVID-19. Although most studies have concluded that school reopening may not pose a significant burden on the epidemic [7, 8], some others have contradicted this statement [9, 10, 11]. Study presented in [9] used an SEIR transmission model to evaluate eight different strategies under varying degrees of mixing by reopening schools for different class groups. They concluded that reopening increases mixing, however this can be constrained to keep the reproductive number under 1 using interventions. A similar conclusion was drawn from a different study [10] that used a stochastic discrete age-structured transmission model that implemented a progressive and prompt reopening strategy with varying percentage of attendance. Study presented in [11] evaluated reopening under two susceptibility assumptions (one where ages below 20 are half as susceptible as adults, and the other where young population is equally as susceptible as adults) and two transmission contexts (high and moderate community transmission). They recommended a hybrid-learning approach within smaller cohorts of 20 students for elementary schools and 10 students for high schools.

## METHODOLOGY

Original version of our AB simulation model for COVID-19 was presented in [12]. The model is particularized for an urban metropolitan region in the U.S. (Miami-Dade County of Florida with 2.8 million population). The AB model generates individual people according to the U.S. census data (by age and occupational distribution), households (by adult and children distribution), schools, workplaces, and community locations. A daily (hour by hour) schedule is assigned to every individual, chosen from a set of alternative schedules, based on their attributes. The model also incorporates temporal changes in the social interventions that were in place during most of 2020.

Interventions include complete and partial lockdowns, school closure and reopening, face mask mandate, and limited contact tracing. The model also considers varying levels of compliances for isolation and quarantine, lower on-site staffing levels of essential work and community places during stay-at-home order, restricted daily schedule of people during various social intervention periods. The AB model reports daily and cumulative values of actual infected, doctor visits, tested, reported cases, hospitalized, recovered, and deaths, for each age category. For more specific details on the input data used for building the model, the algorithmic sequence, and the functional structure of the simulation model, reader is referred to [12]. The model was calibrated using parameters for transmission coefficients at home, work, school, and community places (see [12]). Calibration was done so that the daily cumulative numbers of reported cases from the AB simulation model closely matched the values published in the Florida COVID-19 dashboard until September 30, 2020. Figure 1 shows the daily cumulative reported cases of infection (average from 13 runs with 95% confidence intervals) from AB simulation model. The dotted line in Figure 1 shows the cumulative case growth from the surveillance data from the Florida COVID-19 dashboard for Miami-Dade County [13].

**Figure 1:**
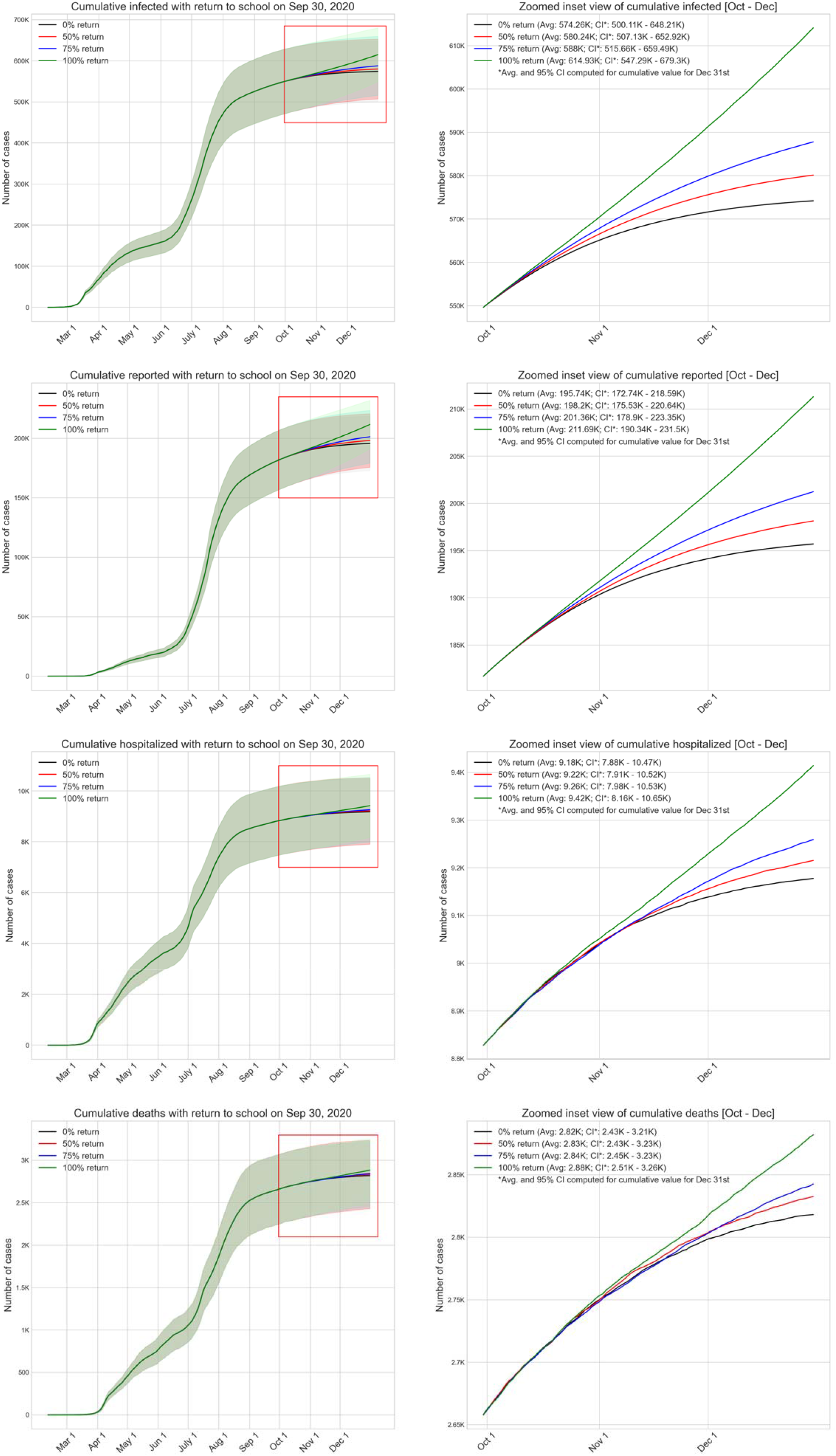
Cumulative plot of the reported cases of infection from AB simulation (average with 95% CI in shade) along with surveillance data in dotted line. {Color}

## RESULTS

We used our model to predict the incremental growth of infected cases, reported cases, hospitalizations, and deaths for the region for various levels of student return and an estimated value of the school transmission coefficient (2.0x). Results are compared with the baseline scenario of 0% return, in which operation of all schools and colleges remains fully virtual until the end of the year 2020. In Miami-Dade County, 21.5% of the population (approximately 600,000 out of 2.8 million people) attend schools (pre-K through 12) and colleges (community colleges, four-year colleges, and universities). Figure 2 summarizes the model outcomes for all four levels of return to school, i.e., 0%, 50%, 75% and 100%. In the model implementation, for scenarios with partial return (50% and 75%), students were rotated. The graphs show average values with 95% C.I. (from thirteen simulation runs with different seeds) of the cumulative numbers of (actual) infected cases, reported cases, hospitalized, and dead. Since the CIs for different level of returns mostly overlap with each other, the average and the CI values for the last day of simulation are noted on the figures. As expected, shortly after the schools and colleges reopen (September 30), the curves begin to diverge. Table 1 summarizes the numerical values of the outcomes from the graphs in Figure 2 for December 31, 2020. The average numbers do increase for reopening with 50% and 75% returns, but the increases are small. For example, the cumulative reported cases by the year end are higher than the base case with 0% return by only 1.26% (p-value 0.59) and 2.87% (p-value 0.22), for 50% and 75% returns, respectively). Notably, the percentage cumulative increase in reported cases for 100% return is almost threefold higher compared to 75% return. When compared with the base case with 0% return, 100% return resulted in a significantly higher cumulative reported cases (by 8.15% with p-value 0.00118). It can also be seen from Figure 2 that for up to 75% return, the daily reported cases reach a decreasing pattern by December 31 and fall near or below a threshold of 100 new daily cases. Whereas for 100% return, the trend for cumulative reported cases remains increasing and the daily reported number on December 31 is still relatively high at 390. It appears that the social mixing process caused by student return of higher than 75% crosses a threshold leading to a sustained (non-decreasing) pattern of new infections until the end of the year. Since the numbers of hospitalization and deaths are derived from the reported cases, graphs for those display similar patterns.

**Figure 2.**
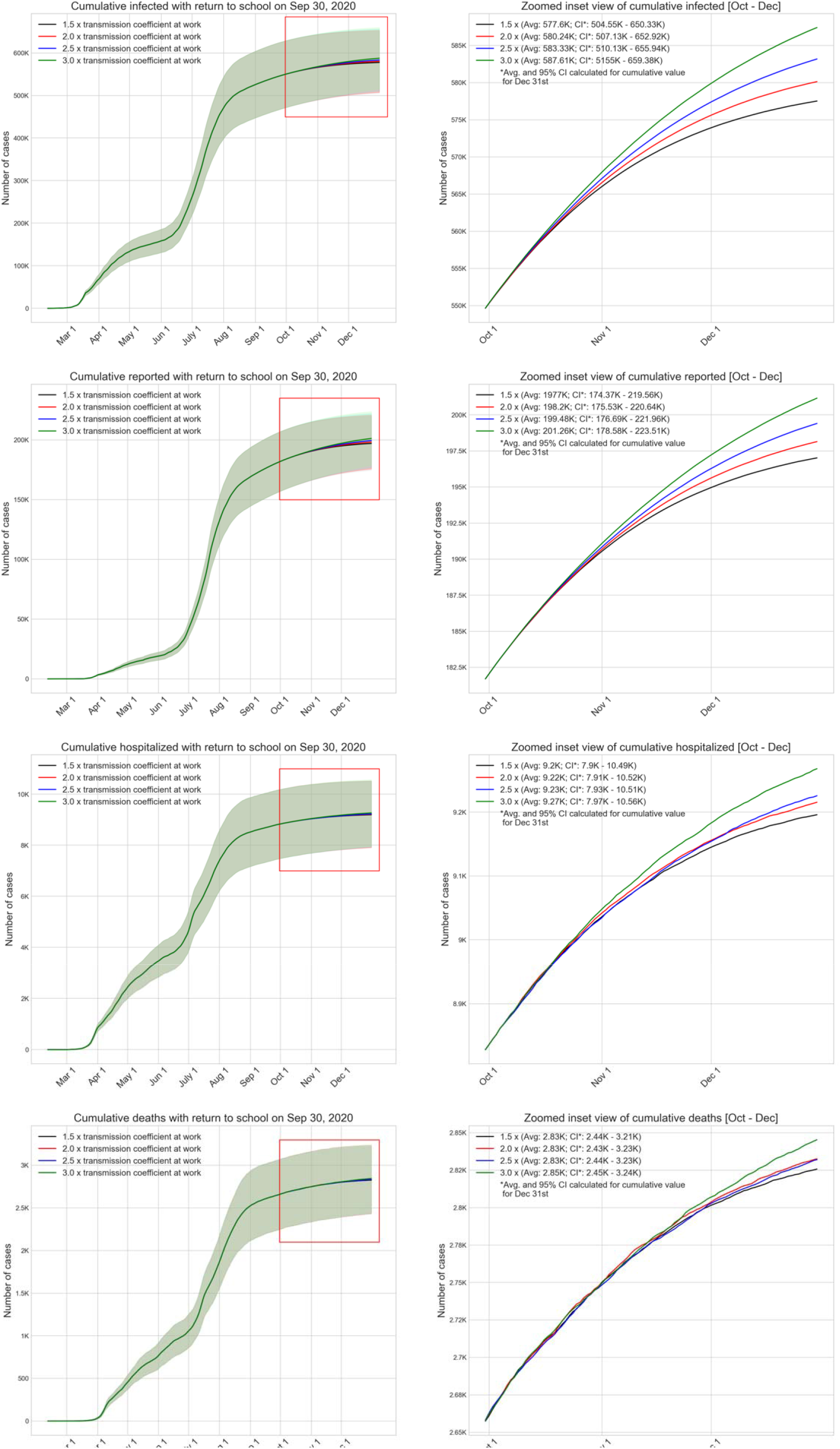
Effect of various levels of school reopening on the cumulative numbers of infected (actual), reported, hospitalized, and dead (with 95% CI in shade) for COVID-19 pandemic in Miami-Dade County, Florida, U.S.A.{Color}

Table 1: Summary of COVID-19 outcomes on Dec 31, 2020 for various levels of student return to school and colleges, as reported by the AB simulation model; the numbers correspond to school transmission coefficient being 2.0 x transmission coefficient at workplaces

Hereafter, we conducted a sensitivity analysis for the transmission coefficient at schools and colleges. Ensuring effective safety protocol at schools and colleges is resource intensive and many school districts don’t have the human and financial wherewithal to adopt a safety protocol (cleaning, sanitizing, social distancing, testing, and quarantining) of the highest standard as recommended by CDC [14]. Consequently, the transmission coefficient may vary significantly among schools and colleges. In our sensitivity analysis, we have examined a number of scenarios where transmission coefficient at schools and colleges are 1.5x, 2.0x, 2.5x, and 3x that of the workplaces (offices and businesses); we have assumed 1.5x as the base case. For this analysis, we maintained the level of student return at 50%, since a survey of Miami-Dade County school district showed that nearly 50% of students intended to return to campus [15].

Table 2: Summary of COVID-19 outcomes on Dec 31, 2020 obtained from the AB simulation model for 50% student return and various transmission coefficient values

Figure 3 depicts the outcomes from scenarios with four different transmission coefficient values at schools and colleges. Table 2 summarizes the numerical values of the outcomes from Figure 3 for December 31, 2020. The impact of increase in transmission coefficient for schools and colleges follow an expected increasing trend. However, the increases in cumulative reported cases for scenarios with 2.0x, 2.5x, and 3.0x transmission coefficients compared to the base case of 1.5x were not statistically significant (p-values are 0.80, 0.59, and 0.36, respectively).

**Figure 3.**
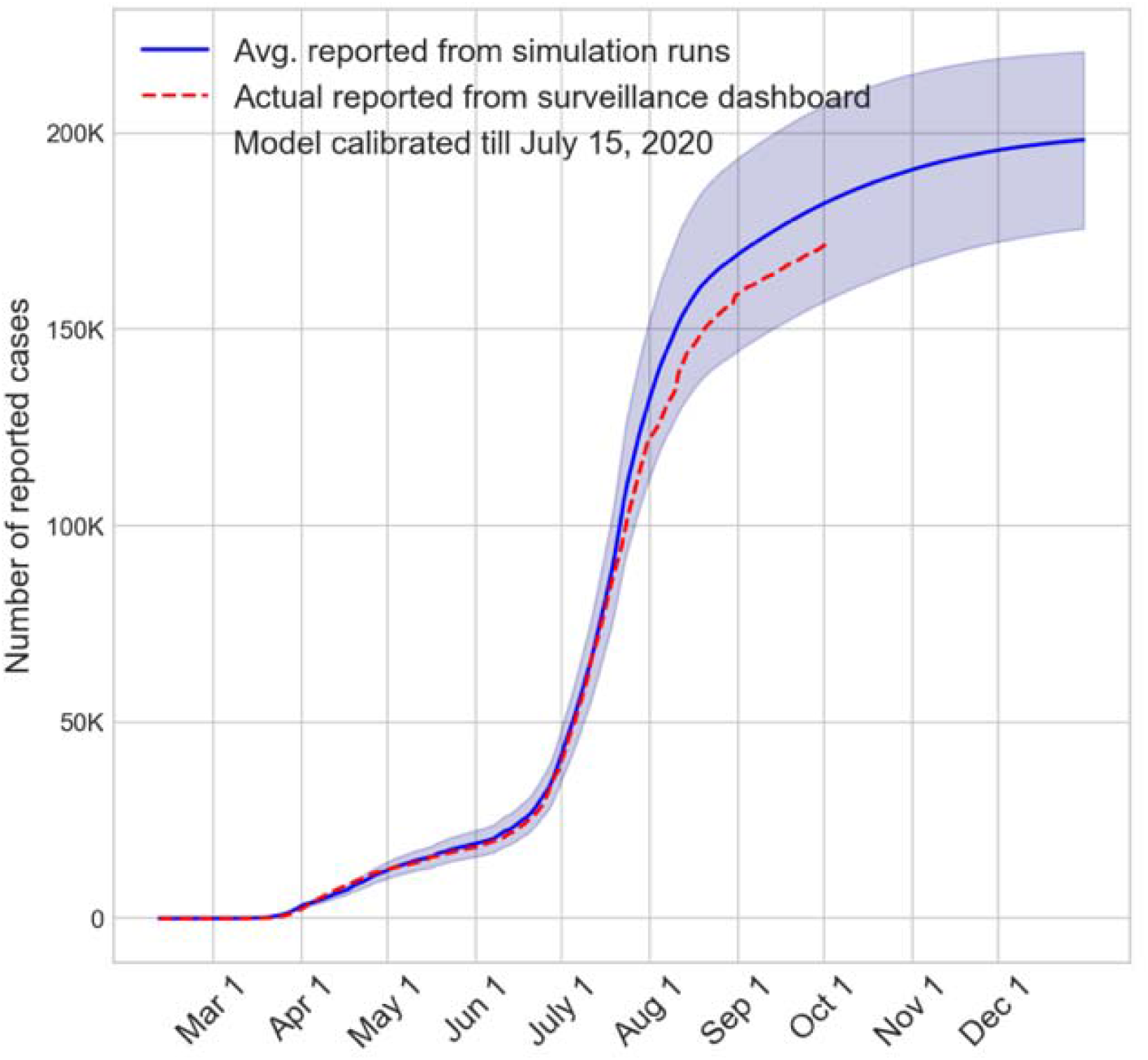
Results of sensitivity analysis for the transmission coefficient parameter characterizing spread of COVID-19 at schools and colleges with 50% of the students returning to campus for in-person instruction {Color}

## DISCUSSION

This study presents an AB simulation model aided investigation of how the reopening of schools and colleges likely has impacted the numbers of actual infected, reported, hospitalized, and dead from COVID-19 pandemic in a densely populated urban region of U.S.A. The AB model is highly flexible and is able to mimic scenarios with a number of chosen levels (%) of students returning to schools and colleges for in-person instruction. The model also accommodates varying levels safety precautions adopted by schools and colleges represented by the values of the transmission coefficient. Results of the model-based investigation can be summarized as follows: 1) for up to 75% return, new additional cases from reopening of schools and colleges is likely to be small, 2) 100% return is expected to cause a steady but modest increase in the number of additional cases till the end of the year, and 3) even when the transmission coefficient at schools and colleges is three times that of the workplaces, a scenario that is indicative of inadequate safety practice, additional number of reported cases is not expected to rise drastically (less than 2.5%). In summary, in order to reduce incremental infections from school reopening, keeping the level of student return below a threshold appears to be important. Also, not having the resources to implement a very high level of campus safety protocol may not be a critical barrier to school reopening. Our findings appear to concur with those reported in the recent studies and media reports [16–21].

The AB model has limitations as it is an abstraction of how a pandemic impacts a large and complex society. A limited number of pre-set daily schedules are used to approximate a highly dynamic contact process, and it does not account for variabilities in types and lengths of interactions. All the schools, work, and community places are assumed to be uniformly distributed in the simulated region, though they usually tend to grow in clusters in urban settings. We also did not implement the quarantining and on-campus testing in schools and colleges. At the time of completing this short communication, we have also completed a study examining two issues for COVID-19: the impact of vaccine (Pfizer/BioNTech and Moderna) administration in the early months of 2021, and an efficacy comparison among different prioritization strategies being considered by various countries and localities. Our findings are presented in [22].

## Data Availability

The datasets used and/or analysed during the current study are available from the corresponding author on reasonable request.

